# Sleep Regularity Index After Stroke: Change Over Time and Its Association with Recovery

**DOI:** 10.64898/2025.12.04.25341669

**Authors:** George D. Fulk, Regina Bell, Keenan Batts, Karen Klingman, Emily Peterson

## Abstract

**Background:** Sleep–wake disruption is common after stroke and may impede recovery. The Sleep Regularity Index (SRI) quantifies the probability that a person is in the same sleep or wake state at the same clock time on consecutive days. This study evaluates SRI during post-stroke recovery, estimating associations with stroke outcomes, and characterizing transitions between SRI states over time.

**Methods:** We analyzed data from 71 participants that were part of a larger on-going study exploring the impact of non-OSA sleep disorders post stroke. Data were collected at 10, 60, and 90 days post stroke. SRI was calculated from sleep-wake data that was collected from wrist worn actigraphy. Covariates included time, National Institutes of Health Stroke Scale (NIHSS), Stroke Impact Scale (SIS), steps/day, sedentary time/day, and Patient Health Questionnaire 9.

Associations between SRI and covariates were estimated using generalized estimating equations. A logistic regression model was used to assess whether clinical variables were associated SRI improvement at 60 days.

**Results:** SRI was low at 10 days post stroke (32.68) and did not significantly change at 60 (34.01, p=0.98) or 90 (38.44, p=0.57) days post stroke. SRI was significantly associated with NIHSS (p=0.04) and SIS (p=0.02). Improvement in SRI was significantly associated with NIHSS (p=0.03).

**Conclusions:** This is the first study that we are aware of to investigate sleep regularity early after stroke. We found that sleep regularity was associated with QOL and stroke severity, and that sleep regularity did not improve after discharge from the hospital.

Clinical Trial Registration: NCT05012605.

---

There is growing evidence that sleep disorders and disturbed sleep negatively impact recovery after stroke. People with stroke and sleep disordered breathing (SDB) are likely to have poorer ability to perform activities of daily living (ADLs) and greater disability than those without SDB.^1^ People with stroke who report poor sleep may also have poorer outcomes.^2^ Disturbances in specific sleep parameters may also negatively influence recovery after stroke. Fleming and colleagues^3^ reported that in people with stroke and acquired brain injury undergoing inpatient rehabilitation between approximately one and three months post stroke/injury, the number of minutes awake after sleep onset (WASO) and sleep fragmentation was associated with poorer motor outcomes and slower recovery during rehabilitation. Ihle-Hansen and colleagues^4^ found that people who slept too little (<7 hours of sleep/night) or too much (>9 hours of sleep/night) during the acute stage after stroke had an increased risk for impaired cognition at three and 18 months after stroke. Bakken and colleagues^5^ reported small associations between WASO and total sleep time (TST) and the ability to perform ADLs during the early stages of recovery after stroke.

However, other studies have reported that specific sleep parameters do not influence recovery after stroke. In people with chronic stroke Fleming and colleagues^6^ found no association between disturbed sleep and motor outcomes. Bakken and colleagues^5^ found no associations between WASO and TST and ADL ability at six months post stroke. Ezeugwu and colleagues^7^ found no association between TST and walking ability, cognition, and physical function at 3 months post stroke.

An important aspect of sleep health that is not captured in sleep parameters that have been commonly explored in people with stroke is regularity, the consistency that a person sleeps across 24 hour periods.^8,9^ Phillips and colleagues^10^ developed the Sleep Regularity Index (SRI) to measure changes in sleep patterns across 24 hour periods with the idea that irregular sleep may cause circadian disruption and thus negatively impact health. The SRI quantifies day-to-day variations in bedtime, wake-up time, and sleep interruptions. Scores range from 0, random sleep-wake patterns with no consistency averaged across 24 hour periods, to 100, perfectly regular sleep-wake patterns averaged across 24 hour periods.

In other populations, the SRI has been associated with and predictive of a variety of key health outcomes. Windred and colleagues^11^ found that greater sleep regularity was associated with a 20%–48% lower risk of all-cause mortality and that sleep regularity was a stronger predictor of all-cause mortality than sleep duration. Lunsford-Avery and colleagues^12^ reported that poorer sleep regularity was associated with and increased 10-year risk of cardiovascular disease, obesity, hypertension, fasting glucose, hemoglobin A1C, and diabetes status. Sansom and colleagues^13^ found that poorer sleep regularity was related to lower physical and mental health-related quality of life in middle aged and older adults.

We are aware of only one study that has examined the association of sleep regularity with stroke outcomes. In a secondary, cross sectional analysis, Schruers and colleagues^14^ found that people with chronic stroke had lower SRI compared to health matched controls. They reported a small association between SRI and depression and a moderate association with self-reported recovery. To the best of our knowledge no studies have examined the SRI longitudinally during the acute and subacute stages of recovery after stroke and its association with stroke outcomes. The purpose of this study was to explore the association between SRI and stroke outcomes at 10, 60, and 90 days post stroke, examine how the SRI may change from 10 to 90 days post stroke and to identify factors that may distinguish between individuals whose SRI improves and those that don’t improve. We also sought to explore the associations between SRI and sleep parameters at 10, 60, and 90 days post stroke, and

## Methods

Participants were recruited from three inpatient rehabilitation hospitals in the Northeast, Southeast, and Midwest of the United States, and are part of a larger, ongoing cohort study to determine the prevalence of non-obstructive sleep apnea (OSA) sleep disorders and their impact on recovery after stroke.^15^ Inclusion criteria were diagnosis of stroke, >= 18 years old, National Institutes of Health Stroke Scale (NIHSS) item 1a score < 2 (alert or not alert but arousable by minor stimulation to obey, answer, or respond), and able to provide informed consent or assent, with the participant’s legal guardian providing consent. Exclusion criteria were pre-stroke diagnosis of OSA, oxygen desaturation index (ODI) >= 15 that was taken at 10 days post stroke, living in nursing home or assisted living prior to stroke, unable to ambulate at least 150’ independently prior to stroke, other neurologic health condition that could impact recovery, pregnancy, recent hemicraniectomy or suboccipital craniectomy, planned discharge > 150 miles from recruiting hospital, and global aphasia as determined by a score of 3 on NIHSS item 9. All participants provided verbal consent or assent (with participant’s legal guardian providing consent), and the study was approved by all local Institutional Review Boards (IRBs) and a central IRB. People with pre-existing OSA or ODI >=15 were excluded because the purpose of the parent study was to determine the prevalence of non-obstructive sleep apnea (OSA) sleep disorders and their impact on recovery after stroke.^15^

When the parent study is completed, data will be made available on request from the authors.

For these analyses, the following stroke related outcomes were collected at 10-, 60-, and 90-days post stroke: Patient Health Questionnaire 9 (PHQ-9),^16^ National Institutes of Health Stroke Scale (NIHSS)^17^ (collected at 10 days post stroke), Stroke Impact Scale (SIS)^18,19^ (collected at 60 and 90 days post stroke). Participants also wore an ActivPal activity monitor (APAM) that measured sitting time/day and number of steps taken/day. The APAM was worn for 5 (at 10 days post stroke) and 7 days (at 60 and 90 days post stroke.) The APAM was wrapped in a nitrile sleeve and secured to the anterior thigh of their least affected lower extremity using a waterproof, transparent film dressing. The APAM is a small (23.5mm wide, 43mm long, only 5mm thick and weighs 9.5 grams [0.93 x 1.7 x 0.2 and weighs 0.34 ounces]) accelerometer-based activity monitor is a valid and reliable method of measuring these behaviors in people with stroke and is recommended by an international consensus group to measure duration in these movement behaviors in the real world^20–24^ and is valid in older adults who are hospitalized.^25^ At 10 days post stroke data was collected while patients were in an inpatient rehabilitation hospital, and at 60 and 90 days post stroke data was taken where the participant was living at the time (home or other location).

To measure sleep time to determine the SRI and other sleep parameters (TST, sleep efficiency [SE], WASO, and number of awakenings [NOA]), participants wore an ActiGraph wGT3X-BT (AG) (https://actigraphcorp.com/actigraph-wgt3x-bt/) 24 hours a day for 5 (at 10 days post stroke) or 7 days (at 60 and 90 days post stroke) on their least affected wrist to estimate sleep parameters.^26^ Actigraphs were set to collect data at a 30Hz sample rate and later uploaded for analysis as 60 second epochs with three-axis motion detection. At the conclusion of each timepoint’s wear period, watch data were transferred from the AG to a study computer for analysis using ActiGraph’s ActiLife v6.13.4 software. Wear times were validated (marked as “wear” if either Troiano method or ActiGraph sensor indicated “wear”), and sleep parameters were determined via the Cole Kripke algorithm and Actigraph sleep period detection method. First-pass software-calculations of time-to-bed and time-out-of-bed were sometimes adjusted prior to calculation of sleep parameter variables, under certain conditions. For overnight timeframes with two separate sleep periods detected by the software, a single sleep period spanning the combination of both originally detected periods was substituted. If the ActiLife software was unable to determine time to bed and time out of bed, every effort was made to manually determine these from activity and light signals in the ActiLife interface. Any sleep periods appearing to be daytime naps were not included in analysis of nightly sleep.

We calculated the SRI directly from the actigraphy-derived sleep–wake time series using the R package sleepreg.^27^ For each participant and timepoint (10, 60, and 90 days post stroke), we first exported minute-by-minute sleep–wake classifications from the ActiGraph data and reformatted them into a 24-hour time series for each calendar day. These data were then imported into R, and the SRI was computed using the package’s implementation of the original Phillips et al. algorithm,^10^ which quantifies the probability that an individual is in the same sleep or wake state at the same clock time on consecutive days. Briefly, for each pair of adjacent days, the algorithm compares the binary sleep–wake state at each minute (or epoch) of the 24-hour cycle; matches are counted as concordant, and the SRI is expressed as a percentage ranging from 0 (completely irregular sleep–wake timing) to 100 (perfectly regular timing). Within each assessment window (10 days post stroke, 60 and 90 days), the package aggregates across all available adjacent-day pairs to yield a single SRI value per participant per timepoint, with higher values indicating more regular sleep–wake patterns. Partial days were excluded and a minimum of 3 full and valid days were required to assess sleep-wake regularity.

### Data Analysis

Because the SIS was only administered at the 60-day and 90-day follow-up visits, participants who contributed data only at the 10-day visit could not contribute valid SIS information for our analyses. To ensure that all participants included in the analytic dataset had the opportunity to contribute SIS scores, we identified individuals with observations exclusively at the 10-day timepoint (i.e., no corresponding 60-day or 90-day measurement) and excluded them from subsequent analyses. This exclusion affected only participants lacking any follow-up beyond the initial assessment (N=23) and did not alter the distribution of covariates among the remaining sample. The final analytic sample included 71 participants.

#### Demographic analysis

Descriptive statistics were used to characterize the sample: age, race, ethnicity, sex, stroke type, stroke location, days from stroke to admission to inpatient rehabilitation hospital, body mass index (BMI), marital status, employment status (60 and 90 days post stroke), living situation (60 and 90 days post stroke), and if receiving rehabilitation services (60 and 90 days post stroke).

#### Longitudinal Associations Between SRI and Stroke Recovery Measures

Time varying covariates measured at all three time points included PHQ-9, daily ambulatory activity (average steps/day), and daily sedentary time (minutes/day). Other covariates were SIS (measured at 60 and 90 days post stroke), and NIHSS (measured at 10 days post stroke).

We conducted all analyses using repeated measures of sleep regularity (SRI) and stroke-related clinical outcomes. Because participants contributed multiple observations over time, we employed Generalized Estimating Equations (GEE) to account for within-person correlation of repeated measures while focusing on population-level associations. We specified population-average GEE models with an identity link and Gaussian family. Our model assessed the association between visit timepoint and SRI, adjusting for standardized clinical and behavioral covariates including NIHSS, PHQ-9, SIS, average steps/day, average sedentary time (minutes/day).

*Model Set-Up:* Let *SRI*_*it*_denote the SRI for participant *i* at visit *t* ∈ (10-days, 60-days, 90-days). Taking 10-days as the reference visit, the model is as follows:

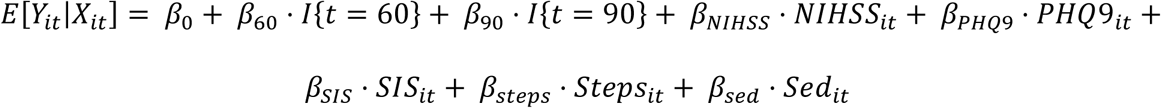

Regression coefficients are interpreted as population-average mean differences in SRI associated with a one-unit increase in each predictor, holding all other variables constant.

#### SRI State Transition Analysis

We examined transitions in SRI states between 10-day and 60-days and evaluated participant characteristics associated with change. To define transition states, we first categorized SRI into Low <33% quantile: SRI <27, Middle (34-66 % quantile: SRI = (28-40)), and High (67+% quantile: SRI=41+) categories based on the observed SRI distribution across time points. For each participant, we compare SRI categories across time points 10-days and 60-days where their sleep patterns plateaued, and each were assigned to one of three ordinal transitions states (Declined, Stable, and Improved) based on this comparison. To construct transition pairs, the data were first ordered by participant identifier and survey timepoint, and consecutive observations were aligned to form (t, t+1) state transitions. Transitions were classified as *Improved* if the participant moved to a higher SRI category at the next timepoint, *Declined* if the next SRI category was lower, and *Stable* if the category remained unchanged. For regression analyses, a binary improvement indicator was created, coded as 1 for transitions in which the SRI state increased and 0 otherwise. We then fit a logistic regression model to assess whether baseline clinical or behavioral factors at time t were associated with the probability of SRI improvement at time t+1. Standardized covariates included: NIHSS, PHQ-9, SIS, average daily step counts, and average daily sedentary time. The model estimated the log-odds of SRI improvement as a function of these predictors, providing effect estimates that describe whether lower stroke severity, greater physical activity, fewer symptoms of depression, or higher cognitive status were associated with increased likelihood of transitioning to a higher SRI category.

To characterize the overall dynamics of SRI changes, we also constructed empirical state-transition matrices. For each SRI category at time t, we tabulated the proportion of participants who transitioned to each of the three states at time t+1, for the 10→60 day interval. These matrices summarize persistence within states as well as the relative frequencies of upward or downward transitions, providing a descriptive complement to the regression analysis. We visualized the transitions graphically, in which flows represented the number of participants moving between SRI states. Together, these analyses characterize both the structural patterns of change in SRI states and the participant-level factors that may predict short-term improvement in sleep regularity during stroke recovery.

#### Associations between SRI and Sleep Parameters

We examined associations between SRI and objective sleep parameters (TST, WASO, SE, and NOA) at each post-stroke time points (10, 60, 90 days). Within each time point, we computed Pearson product-moment correlations between SRI and each sleep parameter, using pairwise complete observations. We report the correlation coefficient *r* for each correlation.

## Results

Data from 71 participants at 10 days post stroke were used in the analysis. At 60 days and 90 days post stroke data from 68 and 59 participants were available. The mean age was 62.68 and 55% were female. 73% of participants had an ischemic stroke and the right hemisphere was the most common location of stroke, Table 2. At 60 and 90 days post stroke the majority lived at home with family or care partner and the most of the participants were still receiving rehabilitation services, Table 3.

**Table 1.** Time-Varying vs. Baseline Covariates Included in GEE Model.

| Variable | Description | Time of Measurement | Time-Varying | Interpretation in Model |
| --- | --- | --- | --- | --- |
| SRI | Day-to-day stability of sleep-wake timing (0-100) | 10, 60, and 90 days | Yes | Outcome varies within individuals over time |
| Time | Study follow-up visit (10, 60, 90 days) | 10, 60, and 90, days | Yes | Adjusts for change over recovery period. |
| SIS | Stroke related quality of life | Measured at 60 and 90 days only | Yes | Reflects evolving quality of life during recovery |
| PHQ-9 | Self-reported depressive symptoms | 10, 60, 90 days | Yes | Captures changes in mood over the recovery period. |
| Average steps per day | Mean daily ambulatory activity | 10, 60, and 90 days | Yes | Reflects shift in mobility and recovery engagement. |
| Average sedentary time per day | Mean daily time sitting and lying | 10, 60, and 90 days | Yes | Reflects shift in sedentary time and recovery. |
| NIHSS | Baseline neurological impairment | 10-days | No (Baseline Only) | Initial stroke severity at baseline. |

\* For participants with a defined SIS score at 60 or 90 days, that value was carried backward to align the participant's 10-day record for model estimation. This ensured SIS represented overall quality of life status rather than implying a true 10-day SIS assessment. Results were robust to sensitivity analyses excluding 10-day rows or omitting carry-forward values.

**Table 2.**
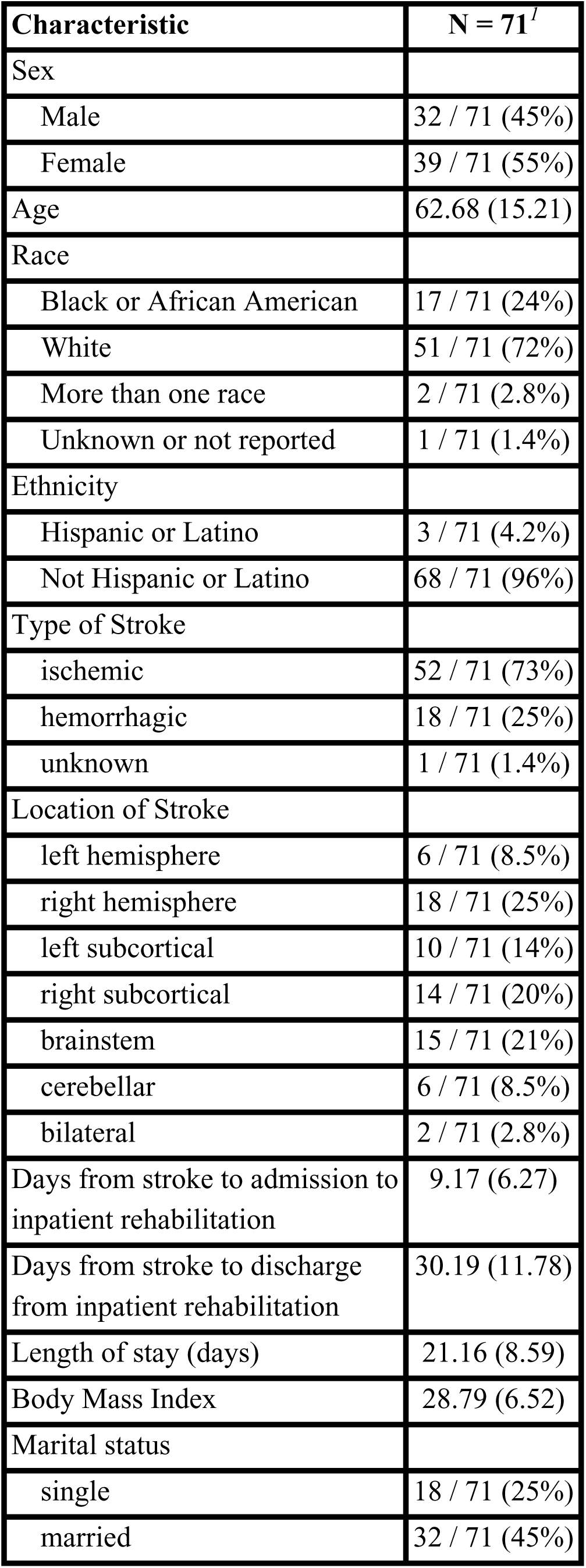
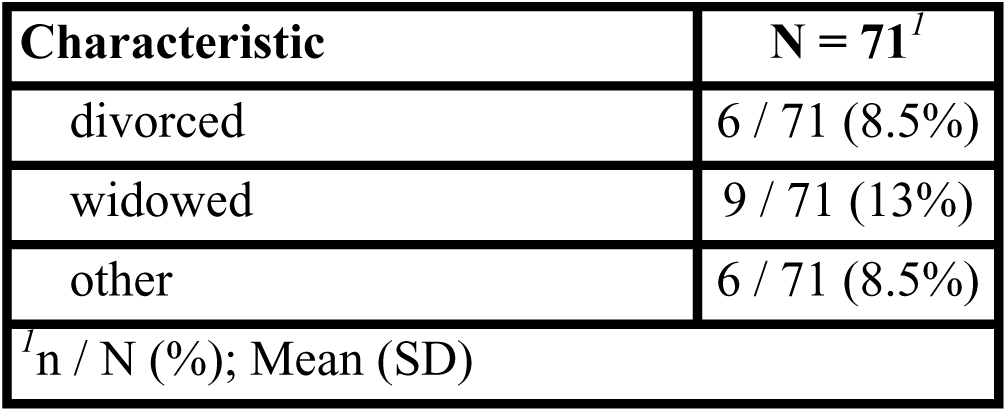
Demographic Data at 10 days post stroke.

| **Characteristic** | **N = 71<sup>I</sup>** |
| --- | --- |
| Sex |  |
| Male | 32 / 71 (45%) |
| Female | 39 / 71 (55%) |
| Age | 62.68 (15.21) |
| Race |  |
| Black or African American | 17 / 71 (24%) |
| White | 51 / 71 (72%) |
| More than one race | 2 / 71 (2.8%) |
| Unknown or not reported | 1 / 71 (1.4%) |
| Ethnicity |  |
| Hispanic or Latino | 3 / 71 (4.2%) |
| Not Hispanic or Latino | 68 / 71 (96%) |
| Type of Stroke |  |
| ischemic | 52 / 71 (73%) |
| hemorrhagic | 18 / 71 (25%) |
| unknown | 1 / 71 (1.4%) |
| Location of Stroke |  |
| left hemisphere | 6 / 71 (8.5%) |
| right hemisphere | 18 / 71 (25%) |
| left subcortical | 10 / 71 (14%) |
| right subcortical | 14 / 71 (20%) |
| brainstem | 15 / 71 (21%) |
| cerebellar | 6 / 71 (8.5%) |
| bilateral | 2 / 71 (2.8%) |
| Days from stroke to admission to inpatient rehabilitation | 9.17 (6.27) |
| Days from stroke to discharge from inpatient rehabilitation | 30.19 (11.78) |
| Length of stay (days) | 21.16 (8.59) |
| Body Mass Index | 28.79 (6.52) |
| Marital status |  |
| single | 18 / 71 (25%) |
| married | 32 / 71 (45%) |

| Characteristic | N = 71<sup>I</sup> |
| --- | --- |
| divorced | 6 / 71 (8.5%) |
| widowed | 9 / 71 (13%) |
| other | 6 / 71 (8.5%) |
| <sup>I</sup>n / N (%); Mean (SD) |  |

**Table 3.** Demographic Data at 60 and 90 days post stroke.

| Characteristic | 60 days N = 68<sup>l</sup> | 90 days N = 59<sup>l</sup> |
| --- | --- | --- |
| Employment |  |  |
| Disabled | 14 / 70 (20%) | 17 / 65 (26%) |
| Full time | 7 / 70 (10%) | 4 / 65 (6.2%) |
| Not employed/unemployed | 7 / 70 (10%) | 6 / 65 (9.2%) |
| Part time | 3 / 70 (4.3%) | 3 / 65 (4.6%) |
| Retired | 31 / 70 (44%) | 32 / 65 (49%) |
| Self-employed | 1 / 70 (1.4%) | 1 / 65 (1.5%) |
| Other | 7 / 70 (10%) | 2 / 65 (3.1%) |
| Receiving Rehabilitation Services | 65 / 70 (93%) | 49 / 65 (75%) |
| Living situation |  |  |
| home with family/caregiver | 56 / 69 (81%) | 56 / 65 (86%) |
| home alone | 5 / 69 (7.2%) | 6 / 65 (9.2%) |
| subacute rehab/skilled nursing facility | 4 / 69 (5.8%) | 2 / 65 (3.1%) |
| other | 4 / 69 (5.8%) | 0 / 65 (0%) |
| Days from stroke to 60 and 90 day interview | 60.39 (4.60) | 93.63 (5.74) |
| <sup>l</sup>n / N (%); Mean (SD) |  |  |

Sleep regularity index was 32.68, 34.01, and 38.44 at 10-, 60-, and 90-days post stroke respectively, Table 4.

**Table 4.**
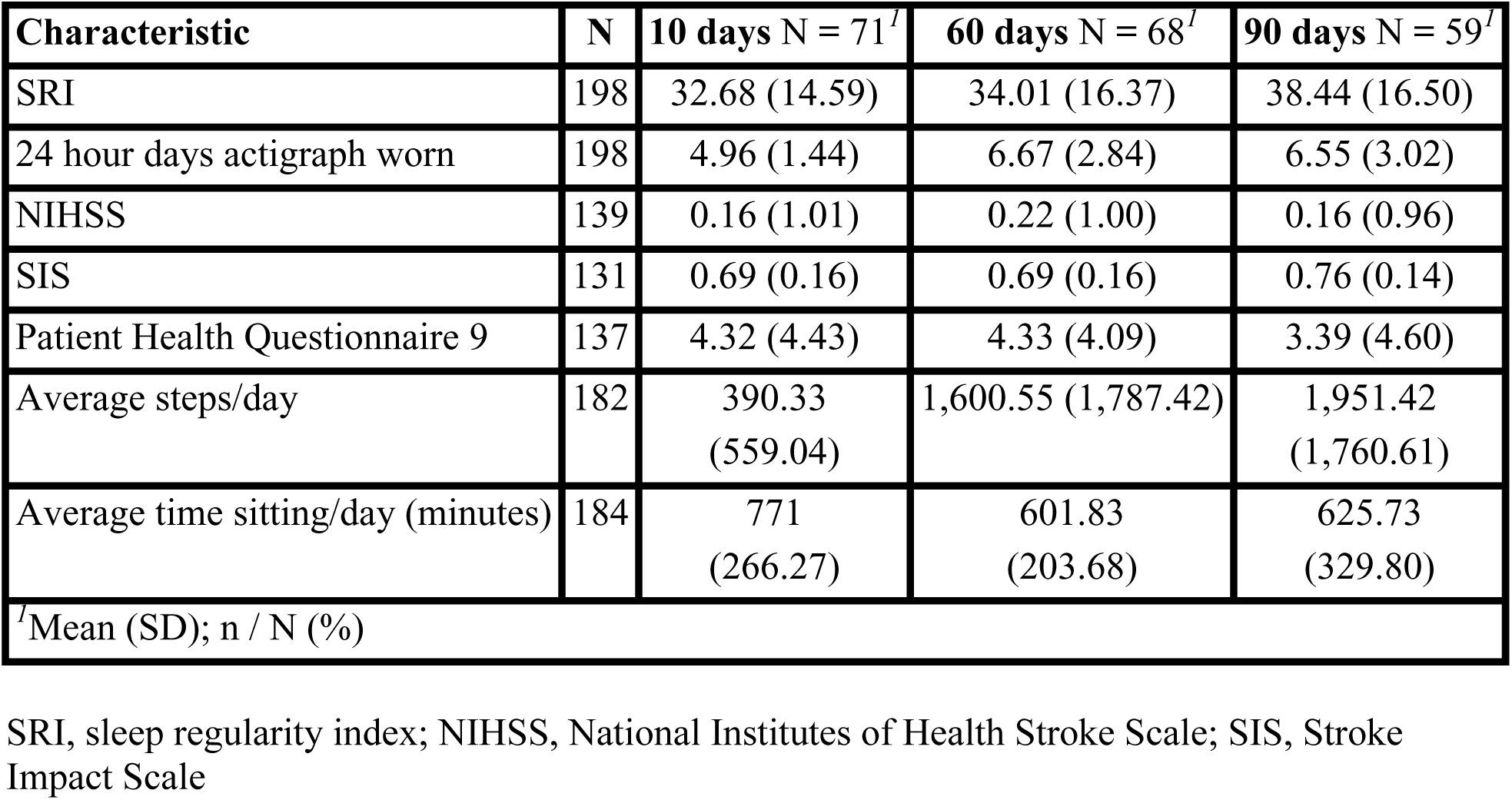
SRI and Stroke Outcomes.

| Characteristic | N | 10 days N = 71<sup>l</sup> | 60 days N = 68<sup>l</sup> | 90 days N = 59<sup>l</sup> |
| --- | --- | --- | --- | --- |
| SRI | 198 | 32.68 (14.59) | 34.01 (16.37) | 38.44 (16.50) |
| 24 hour days actigraph worn | 198 | 4.96 (1.44) | 6.67 (2.84) | 6.55 (3.02) |
| NIHSS | 139 | 0.16 (1.01) | 0.22 (1.00) | 0.16 (0.96) |
| SIS | 131 | 0.69 (0.16) | 0.69 (0.16) | 0.76 (0.14) |
| Patient Health Questionnaire 9 | 137 | 4.32 (4.43) | 4.33 (4.09) | 3.39 (4.60) |
| Average steps/day | 182 | 390.33<br>(559.04) | 1,600.55 (1,787.42) | 1,951.42<br>(1,760.61) |
| Average time sitting/day (minutes) | 184 | 771<br>(266.27) | 601.83<br>(203.68) | 625.73<br>(329.80) |
| <sup>l</sup>Mean (SD); n / N (%) |  |  |  |  |

SRI, sleep regularity index; NIHSS, National Institutes of Health Stroke Scale; SIS, Stroke Impact Scale

### Longitudinal Associations Between SRI and Stroke Recovery Measures

In the adjusted population-average GEE model, SRI did not differ significantly across follow-up timepoints. Compared with the 10-day visit, mean SRI was not significantly different at 60 days (β = 0.10, SE = 4.53, p = 0.98) or 90 days (β = 2.80, SE = 4.90, p = 0.57). Baseline stroke severity (NIHSS) showed a positive but non-significant association with SRI (β = 2.86, SE = 1.99, p = 0.15). Depressive symptoms (PHQ-9), daily ambulatory activity (steps/day), and sedentary time were also not significantly associated with SRI in adjusted models (all p > 0.40). Self-reported QOL (SIS) demonstrated the largest effect estimate and a trend toward higher sleep regularity ( β = 28.76, SE = 15.70, p = 0.067). The working correlation estimate (α = 0.37) indicated moderate within-participant correlation in repeated SRI measurements. Overall, the model suggests that greater QOL may correspond to more regular sleep–wake patterns during post-stroke rehabilitation, although the association did not reach conventional statistical significance in this sample.

### Follow-Up Analysis Excluding Time

In a secondary model that removed the visit timepoint variable, baseline stroke severity (NIHSS) was significantly associated with lower sleep regularity (β = 3.71, SE = 1.80, p = 0.04). SIS was also significantly associated with higher SRI (β = 32.73, SE = 13.98, p = 0.02), while depressive symptoms, steps per day, and sedentary time remained non-significant, Table 5. The emergence of a significant NIHSS effect in this model reflects the fact that the overall improvement in sleep regularity over time is partially shared with baseline severity; once time is omitted, that recovery-related variation is absorbed by NIHSS. When time is included, this shared trend is accounted for directly, reducing NIHSS to a non-significant predictor in the primary model.

**Table 5.** Generalized Estimating Equation.

| Independent variables | Estimate | Standard Error | Wald | p |
| --- | --- | --- | --- | --- |
| Intercept | 9.434962 | 11.533002 | 0.67 | 0.413 |
| NIHSS | 3.705580 | 1.800692 | 4.23 | 0.040* |
| PHQ-9 | -0.250047 | 0.310487 | 0.65 | 0.421 |
| SIS | 32.728295 | 13.981544 | 5.48 | 0.019* |
| Steps/day | 0.000424 | 0.001014 | 0.17 | 0.676 |
| Sitting time | 0.001842 | 0.006172 | 0.09 | 0.765 |

\*p<0.05
NIHSS, National Institutes of Health Stroke Scale; PHQ-9, Patient Health Questionnaire; SIS, Stroke Impact Scale

### SRI State Transition Results

Out of the 71 participants, 22 (31%) showed a decline in SRI state, 22 (31%) showed an improvement, and 27 (38%) showed a stable unchanged SRI state. Results show improvement in SRI transitions states was solely significantly associated with NIHSS (p-value = 0.03) when controlling for all other predictive factors. The odds ratio of having an improved state was 2.07 (95% CIs = (1.05, 4.12)). This indicates that as the initial stroke severity worsened (higher NIHSS score) the odds of improvement from 10 to 60 days increased.

To clarify why higher NIHSS (greater stroke severity) was associated with greater odds of SRI improvement, we conducted exploratory analyses. At 10 days, NIHSS was weakly but negatively related to baseline SRI (β = −0.15, p = 0.21), indicating that participants with more severe stroke began with somewhat lower SRI. Baseline SRI category was the strongest determinant of improvement from 10 to 60 days, with 54% of participants in the Low category improving, compared with 31% in Mid. When baseline SRI category was included in a penalized logistic regression model to address separation, NIHSS was no longer associated with improvement, indicating that the apparent effect of NIHSS reflected the greater “room to improve” among individuals starting with very low sleep regularity. Figure 1 displays the distribution of NIHSS scores across baseline SRI categories showing that participants in the Low SRI group generally had higher stroke severity than those in the Mid or High groups, i.e., more room for improvement at 60/90 days.

**Figure 1.**
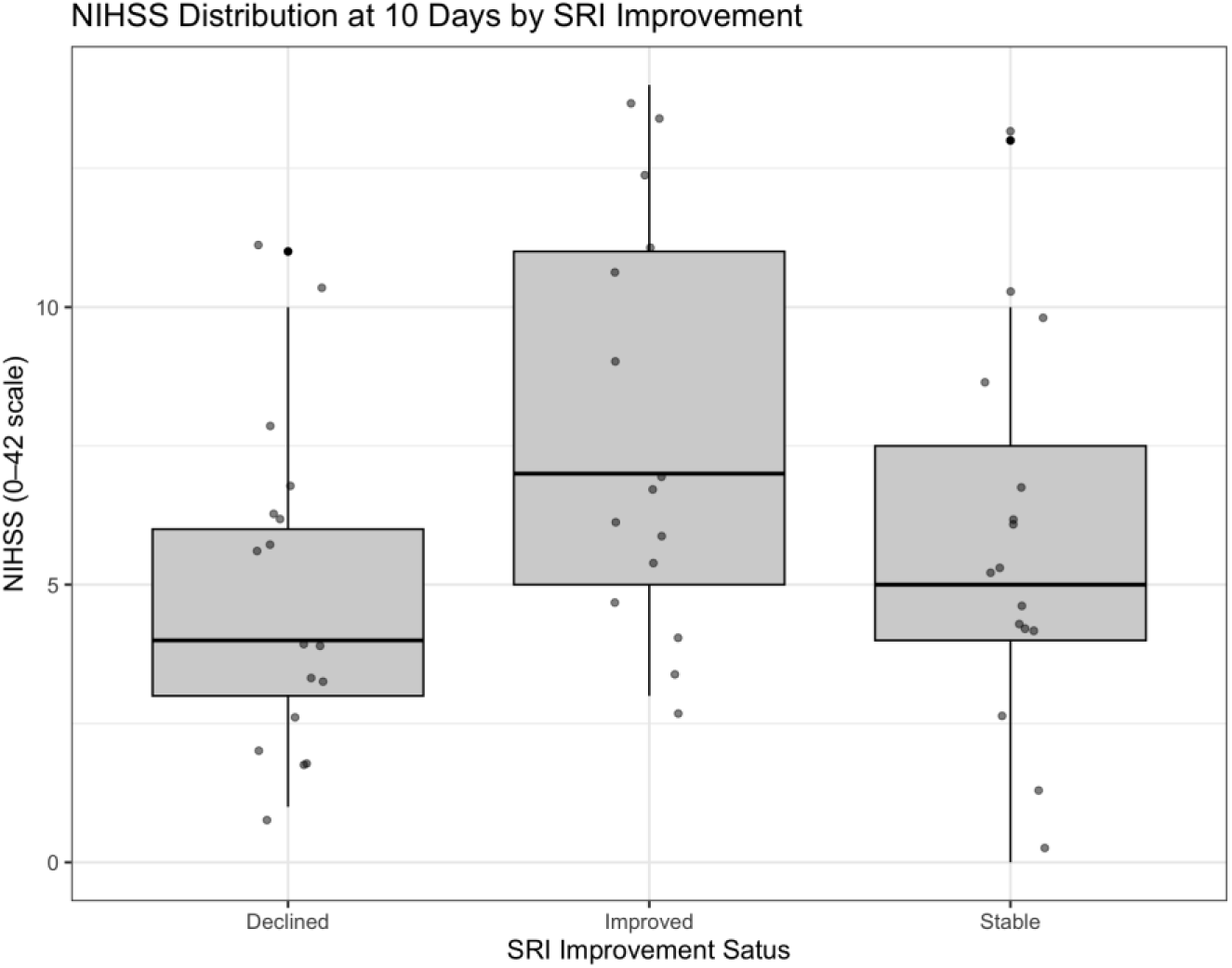
Distribution of NIHSS scores across baseline Sleep Regularity Index (SRI) categories at 10 days post stroke.

Figure 2. shows the breakdown of transition states from 10 to 60 days in which the percentage of each transition is calculated as the number of participants in SRI state X at 60-days out of the number of people in SRI state Z at 10 days. For example, out of the participants in Low SRI state at 10 days, 52.4% remained in the Low SRI group at 60 days. The Figure 2. illustrates that if participants started in the Low or High SRI states, the majority remained stable across times. For participants that started in the Mid SRI state, the breakdown of transition to Low, Mid, High was split evenly. It is important to note, that SRI was generally low across the entire population, where on average health individuals are expected to have an SRI of 80% or above.^27^

**Figure 2.**
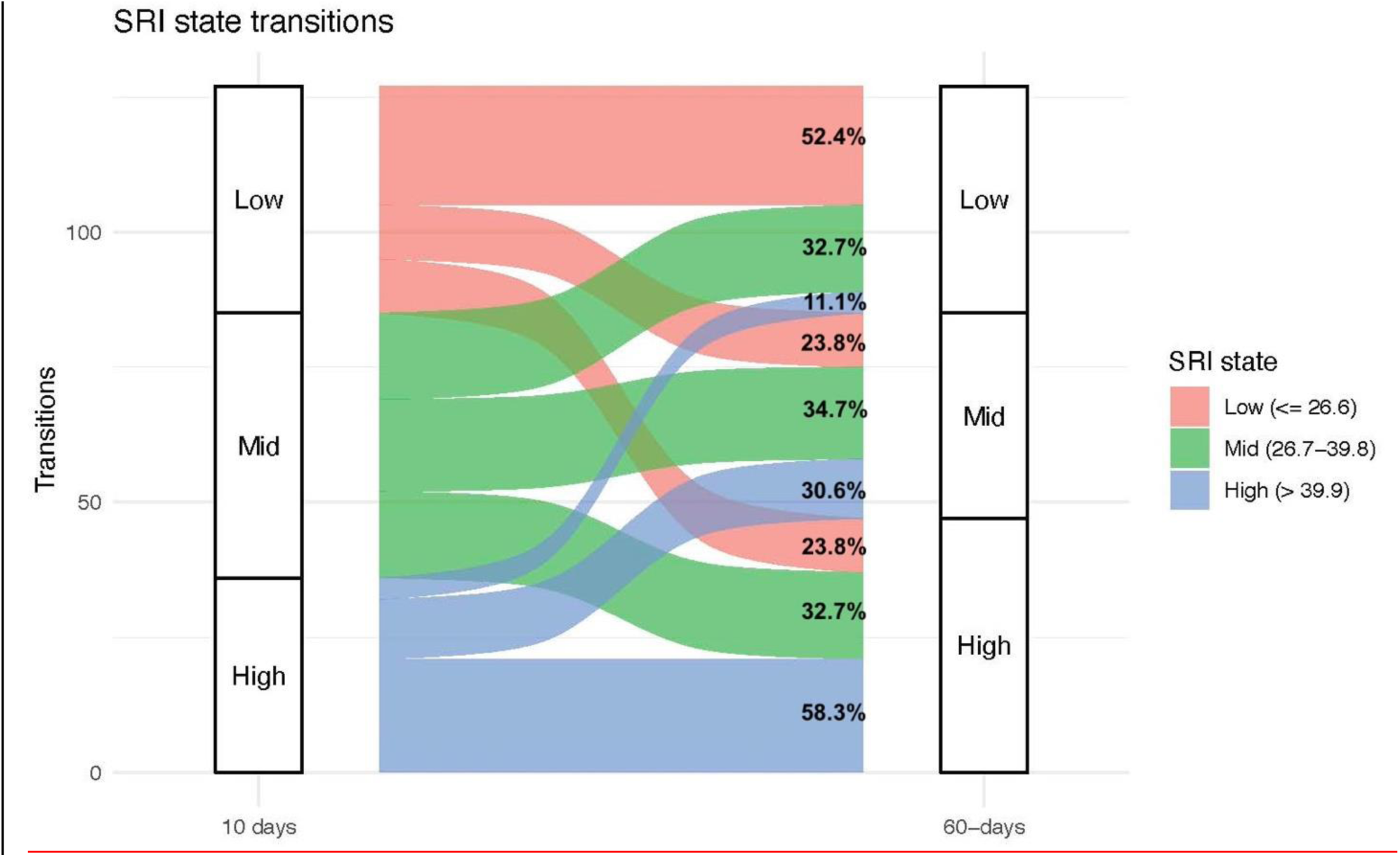
SRI State Transitions.

### Correlations between SRI and Sleep Parameters

There were small to moderately significant associations between SRI and sleep parameters at 10 and 60 days post stroke, but not at 90 days post stroke. At 10 days post stroke SRI was associated with SE (r=0.34), WASO (r=-0.32), and NOA (r=-0.25). At 60 days post stroke SRI was associated with SE (r=0.39) and WASO (r= – 0.40), see Figures 3a-c and table 6.

**Figure 3a.**
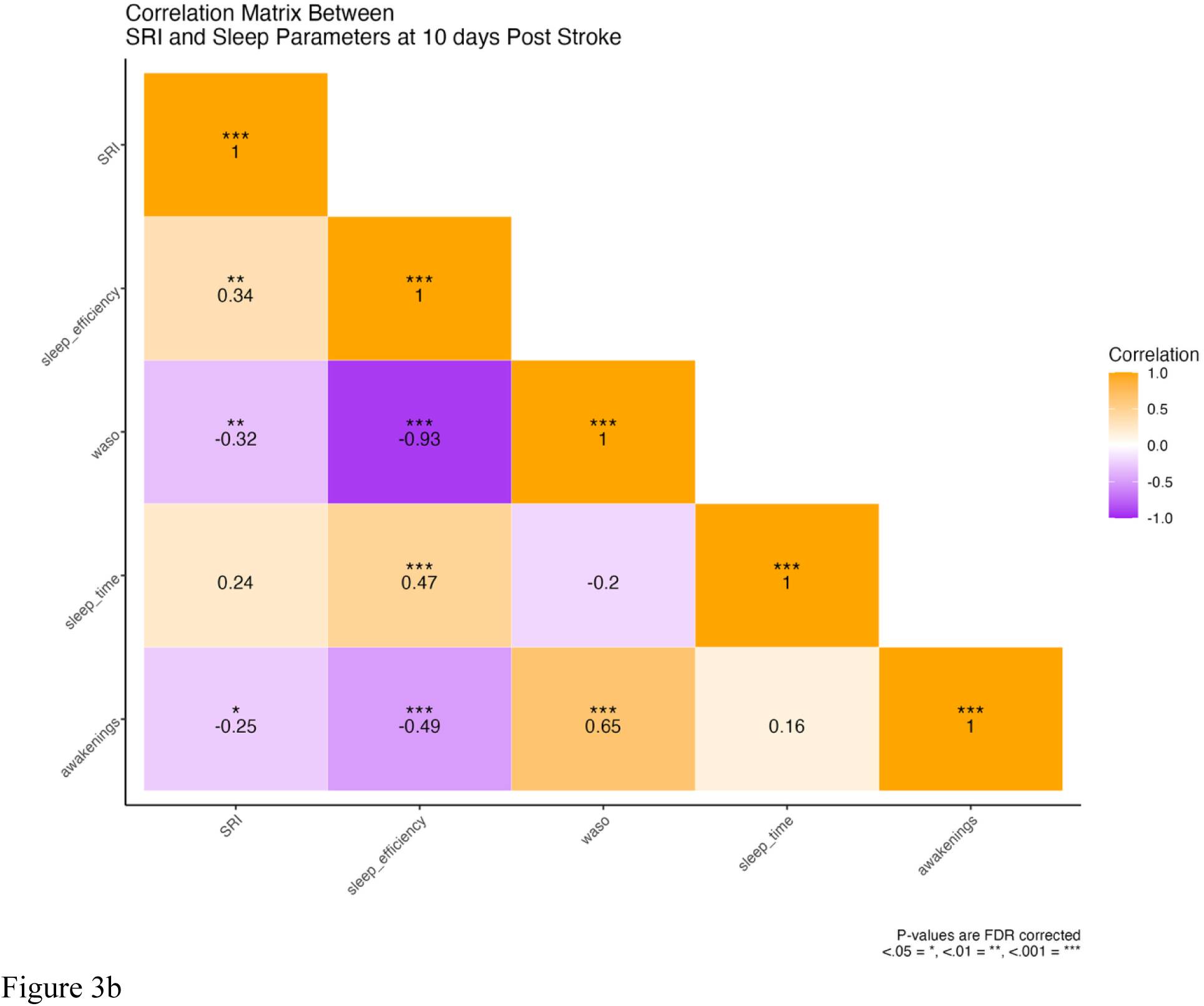
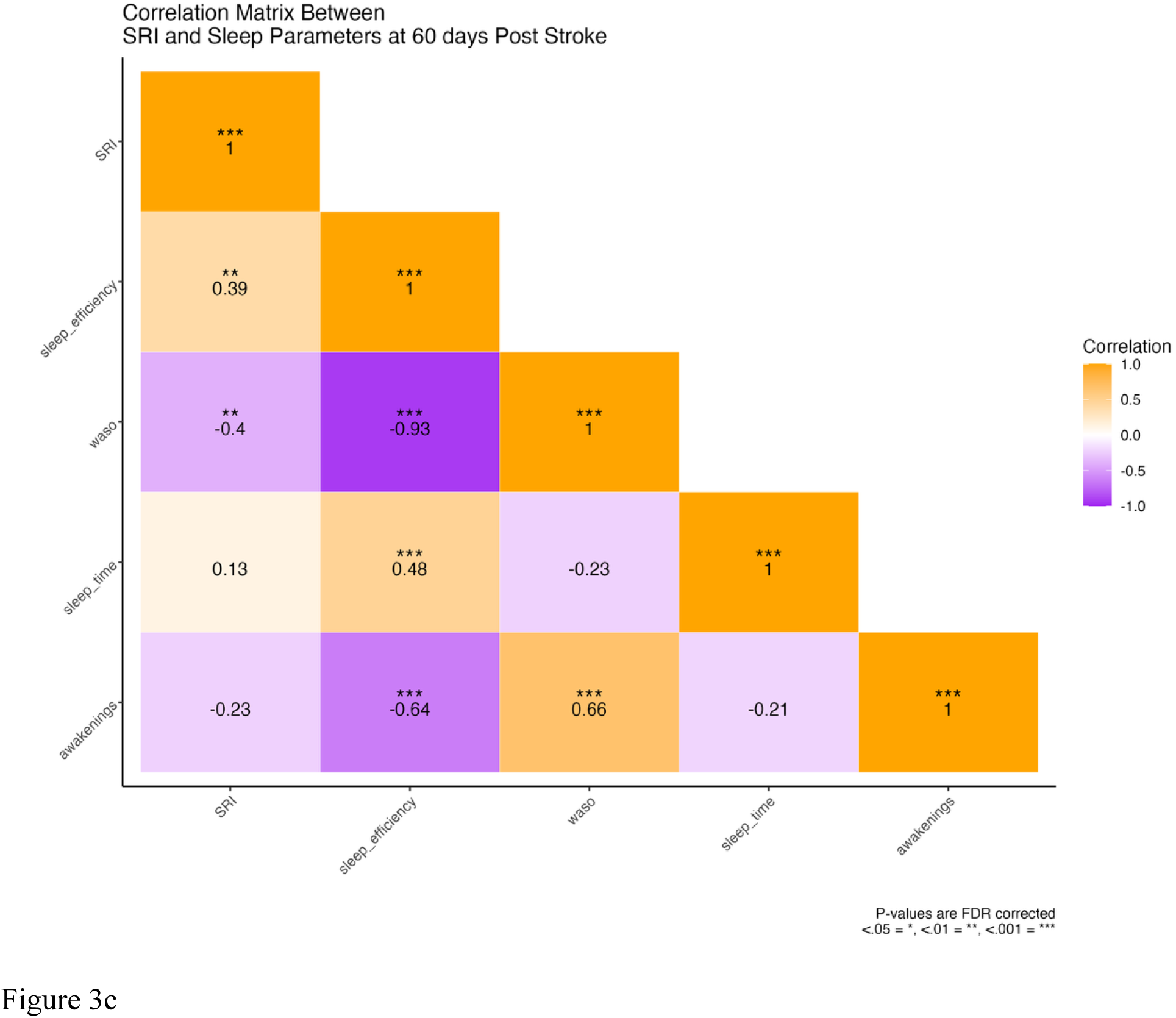
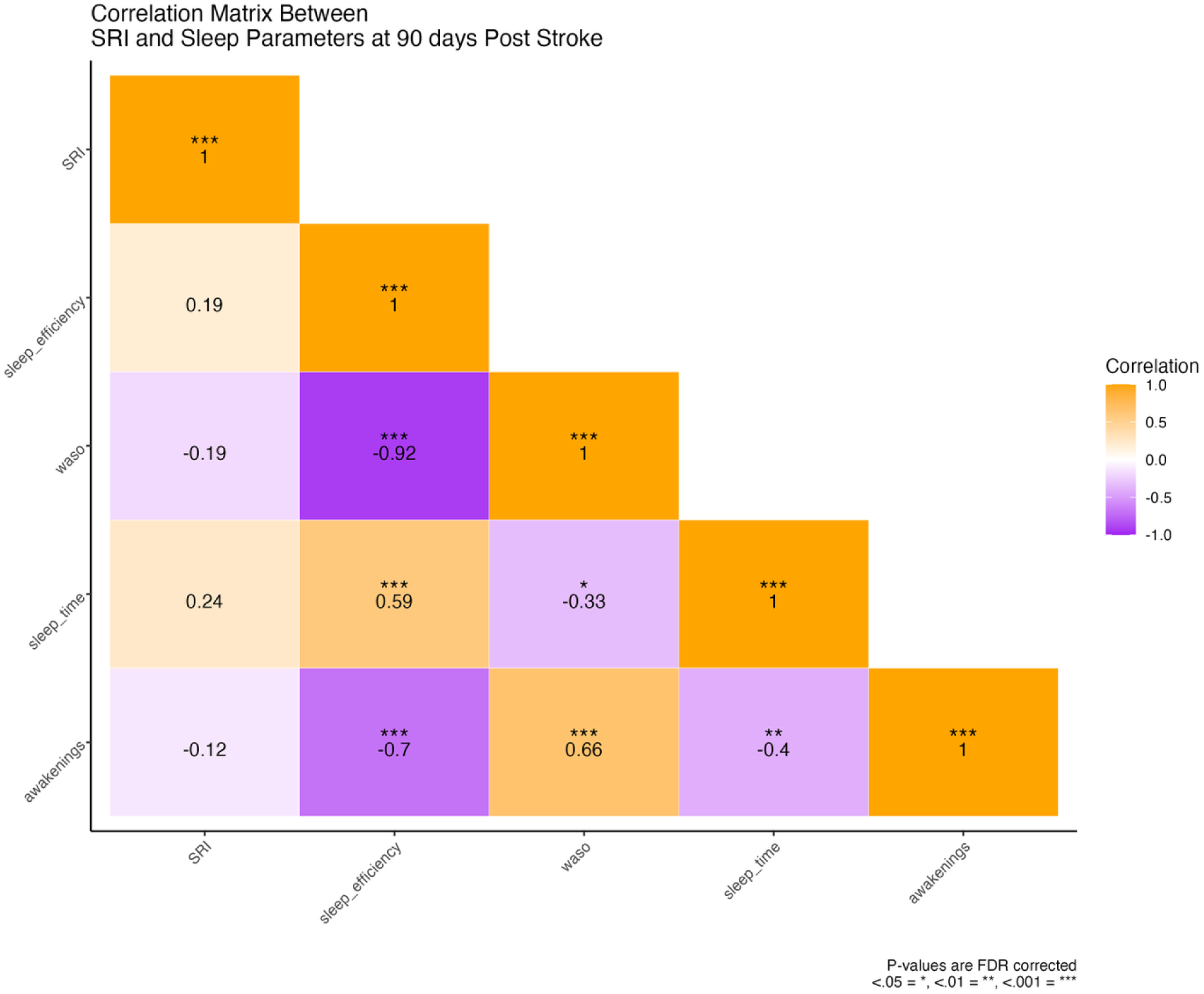
Association Between SRI and Sleep Parameters.

**Table 6.** SRI and Sleep Parameters.

| Characteristic | 10 days N = 71<sup>1</sup> | 60 days N = 68<sup>1</sup> | 90 days N = 59<sup>1</sup> |
| --- | --- | --- | --- |
| SRI | 32.68 (14.59) | 34.01 (16.37) | 38.44 (16.50) |
| Sleep Efficiency | 84.53 (7.92) | 81.89 (8.66) | 81.51 (9.04) |
| WASO | 75.24 (41.10) | 96.11 (54.64) | 99.58 (61.18) |
| Sleep Duration (minutes) | 409.26 (104.97) | 456.75 (92.17) | 457.53 (93.90) |
| Awakenings | 12.74 (5.54) | 16.63 (6.99) | 16.35 (7.00) |
| <sup>1</sup>Mean (SD); n / N (%) |  |  |  |

SRI, sleep regularity index; WASO, wake after sleep onset

## Discussion

The impact of sleep on recovery after stroke is complex and only beginning to be explored. This is the first study to explore sleep regularity early after stroke and how it evolves during the first three months after stroke. In this cohort of people with stroke without OSA, sleep was highly irregular early and did not significantly change at 3 months post stroke. More regular sleep was associated with higher stroke related quality of life at 60 and 90 days post stroke and greater stroke severity at 10 days post stroke. Individuals with greater stroke severity initially were more likely to demonstrate improvements in sleep regularity at 60 days. Sleep regularity was also associated with some sleep parameters early after stroke but not at 90 days post stroke.

There is a growing body of literature on the importance of sleep regularity for health,^28^ and it is linked to a variety of poor health outcomes such as cardiovascular disease, metabolic dysfunction, depression, and mortality.^11,13,29–31^ Sleep was highly irregular in our cohort of people with stroke without OSA compared to available population normative values. Using data from over 60,000 people in the UK Biobank, Windred and colleagues^27^ reported a median SRI of 81.0 with an interquartile range of 73.8-86.3. 99% subjects in the sample had an SRI between 36.0 and 95.0.

The SRI of the participants in our study was similar, although slightly more irregular, compared to people with chronic stroke reported by Schruers and colleagues.^14^ In a secondary, cross sectional analysis, Schruers and colleagues^14^ found that people five years post stroke had a mean (± sd) of 41.21 (12.58), while in our sample at 10, 60, and 90 days post stroke the mean (±sd) SRI was 32.68 (14.59), 34.01 (16.37), and 38.44 (16.50) respectively. The extent that irregular sleep contributes to poor outcomes after stroke should be further explored. The SRI may be a useful way to assess sleep and its impact on recovery after stroke.

Our findings that sleep regularity was associated with stroke related quality of life (SIS) and stroke severity (NIHSS) provides further support to the growing body of evidence of the importance of sleep on recovery after stroke. Schruers and colleagues^14^ found a moderate association between SRI and self-reported quality of life in people with chronic stroke. Fulk and colleagues^2^ found that self-reported quality of life was lower in people that reported sleep problems negatively impacted their function compared to those that reported no sleep problems. Others have found that specific abnormal sleep parameters such as sleep efficiency, total sleep time, and WASO are associated with poorer outcomes after stroke.^3–5^ Irregular sleep may interfere with patients ability to fully participate in rehabilitation interventions after stroke, thereby impacting their recovery.

There is a complex, bi-directional relationship between sleep and depression.^32^ Depression is common after stroke^33,34^ with multiple neurological and psychosocial contributors to its prevalence.^35^ It was somewhat surprising that mood (PHQ-9) was not associated with SRI. A few other studies have reported an association between mood/depression and sleep disturbances in people with stroke. Schruers and colleagues^14^ reported a small association between SRI and mood in people with chronic stroke. Davis and colleagues,^36^ Tenorio and colleagues,^37^ and Fan and colleagues^38^ all have reported that people with self-reported poor sleep after stroke are more likely to have increased levels of depression. The SIS contains a section with nine questions on mood/emotions. It is possible that this section of the SIS partially contributes to the association between the total SIS score and sleep regularity reducing PHQ-9 to a non-significant predictor in the model The relationship between irregular sleep and depression after stroke may also be bidirectional. Further research is needed to explore the potential benefits of improving sleep during recovery after stroke to improve mood.

Most studies examining the impact of sleep on recovery after stroke have primarily focused on self-report of sleep duration and quality of sleep with little attention to sleep regularity. We are aware of only one other study that has explored sleep regularity and its association with recovery after stroke, and this was done cross sectionally in people with chronic stroke. It was unexpected that sleep regularity did not improve over the first 90 days post stroke. We thought that once participants were out of the hospital and back in their home environment, they would be able to establish a more regular sleeping pattern. However, this was not the case. It may be that during this still relatively early stage of recovery it is challenging for people with stroke to maintain a regular schedule as they adjust to a new life. Another possibility is that irregular sleep patterns are entrained while in the hospital, making it difficult for people with stroke to adopt a more regular sleep pattern once out of the hospital. Most guidelines to promote healthy sleep focus on duration^39^ and quality,^40^ it may also be important to promote a regular sleep pattern after stroke to enhance recovery and to address this early in recovery.

We found small associations between SRI and sleep parameters (SE, WASO, and NOA) at 10 and 60 days post stroke, but none at 90 days post stroke. This is consistent with findings from other studies. Schruers and colleagues^14^ found a small association between SRI and TST in people with chronic stroke, but no associations with other sleep parameters.

A limitation of our study is that we did not include people with stroke who have OSA, so that our findings cannot be generalized to all people with stroke. Community dwelling, neurologically intact individuals with greater sleep irregularity are more likely to have OSA.^41^ It is possible that people with stroke and OSA also have irregular sleep. However, we are not aware of any studies that have examined sleep regularity in people with stroke and OSA. Importantly, we found that people with stroke without OSA have irregular sleep, so it is important to assess sleep in people with stroke without OSA.

Although we found that irregular sleep was associated with poorer QOL and greater initial stroke severity, we cannot determine why some of the participants developed irregular sleep patterns and others did not. Future research should explore the causal links between stroke and irregular sleep so that effective interventions to improve sleep regularity can be developed.

## Data Availability

Data will be made available on request from the authors when the parent study is completed.

## Conclusions

There is a growing understanding of the importance of sleep on recovery after stroke. The sleep regularity index may be a useful method to assess sleep over time as it provides unique insights into sleep/wake patterns across a 24 hour period. This is the first study that we are aware of to investigate sleep regularity early after stroke and to explore how it evolves during the first three months post stroke. We found that sleep regularity was associated with QOL and stroke severity, and that sleep regularity did not improve upon discharge from the hospital. Improving sleep regularity may help enhance QOL post stroke, but further research is needed to develop effective interventions.

## Disclosure

The authors have no conflicts of interest related to this manuscript to disclose. Funding: This research was supported by the NIH NINR, Award Number R01NR018979.

## Non-standard Abbreviations and Acronyms

SDB: sleep disordered breathing
ADLs: activities of daily living
WASO: awake after sleep onset
TST: total sleep time
SRI: Sleep Regularity Index
OSA: obstructive sleep apnea
NIHSS: National Institutes of Health Stroke Scale
ODI: oxygen desaturation index
IRBs: Institutional Review Boards
PHQ-9: Patient Health Questionnaire 9
NIHSS: National Institutes of Health Stroke Scale
SIS: Stroke Impact Scale
AG: ActiGraph
APAM: ActivPal activity monitor
SE: sleep efficiency
NOA: number of awakenings
BMI: body mass index
GEE: Generalized Estimating Equations

## Notes

### Competing Interest Statement

The authors have declared no competing interest.

### Clinical Trial

Clinical Trial Registration: NCT05012605.

### Author Declarations

WCG IRB (20202548). SUNY Upstate Medical University IRB (1643499) University of Kansas IRB (00148016). Emory University: 00007229

